# HUMAN FECAL BILE ACID ANALYSIS AFTER INVESTIGATIONAL MICROBIOTA-BASED LIVE BIOTHERAPEUTIC DELIVERY FOR RECURRENT *CLOSTRIDIOIDES DIFFICILE* INFECTION

**DOI:** 10.1101/2022.08.30.22278604

**Authors:** Romeo Papazyan, Nicky Ferdyan, Karthik Srinivasan, Carlos Gonzalez, Bill Shannon, Ken Blount, Bryan C. Fuchs

**Affiliations:** Ferring Research Institute, San Diego, CA, USA; BioRankings LLC, St. Louis, MO, USA; Rebiotix Inc., a Ferring Company, Roseville, MN, USA

## Abstract

Intestinal microbiome disruption is associated with recurrent *Clostridioides difficile* infection (rCDI), which poses a high risk of morbidity and mortality. Microbiome-based therapeutics are increasingly evaluated as a strategy to reduce rCDI, and their proposed mechanisms include restoration of the microbiota and microbiota-mediated functions, including bile acid (BA) metabolism. This study reports the development of a highly quantitative and sensitive assay for targeted metabolomic assessment of bile acids, and the application of the assay to profile bile acid composition in a Phase 2 trial of the investigational microbiota-based live biotherapeutic RBX2660 for reduction of rCDI (PUNCH CD2; NCT02299570). Participants were asked to provide stool samples before and up to 24 months after treatment. A liquid chromatography tandem mass spectrometry method was developed to extract and quantify 35 BAs from a total of 113 participant stool samples from 27 rCDI participants treated with RBX2660 in the double-blinded, placebo-controlled clinical trial. The results demonstrate a high-confidence assay as represented by sensitivity, linearity, accuracy, and precision of the output measurements of BAs. When the assay was utilized to assess stool samples from the clinical trial participants, primary BAs were the dominant BA species at baseline, consistent with the expected loss of commensals after broad-spectrum antibiotic treatment. As early as 7 days after RBX2660 administration, there was a significant drop in primary BAs concurrent with increased secondary BAs, and this profile was sustained through 24 months after RBX2660 administration. Taken together, we describe a robust assay that demonstrates altered BA metabolism associated with RBX2660 administration, shifting towards a profile that is consistent with a healthier BA profile and clinical response.

## INTRODUCTION

Recurrent *Clostridioides difficile* infection (rCDI) is a significant public health concern with insufficiently effective treatment options. While antibiotic treatment remains the standard of care for rCDI, antibiotics fail to rectify the root cause of CDI recurrence—disruption of healthy gut microbiota, known as microbiome dysbiosis—and recurrence after antibiotics remains high. rCDI-associated dysbiosis is characterized by significant changes in the composition and diversity of bacterial microbiota. Consequently, each successive CDI recurrence after antibiotic treatment increases the chance for subsequent recurrence(1).

Alteration of the fecal composition of bile acids (BAs) is one of the functional mechanisms by which dysbiosis may promote CDI recurrence(2,3). The primary BAs cholic acid (CA) and chenodeoxycholic acid (CDCA) are synthesized and conjugated with taurine or glycine in the liver, which are then released into the intestinal tract via the gallbladder. Healthy commensal bacteria deconjugate and chemically modify primary BAs to a diverse set of secondary BAs(4). Lithocholic acid (LCA) and deoxycholic acid (DCA) are secondary BAs that dominate the healthy fecal BA pool and are derived from primary BA through dehydroxylation reactions of the 7-position carbon carried out by commensal bacteria(4,5). Secondary BAs contribute to many homeostatic processes in host physiology including resistance to infection by pathogens like *C. difficile*(4). In rCDI patients, dysbiosis results in an accumulation of primary BAs in the intestinal tract, some of which can promote *C. difficile* spore germination and vegetative growth(4,6). Thus, the balance of BA compositions is thought to be a key determinant of resistance or susceptibility to rCDI.

To address the dysbiosis in rCDI patients and thereby reduce recurrence, several investigational microbiota-based therapeutics are in clinical development, including RBX2660— a stabilized, microbiota-based investigational live biotherapeutic(7). In several clinical trials, RBX2660 reduced CDI recurrence and normalized trial participants’ microbiota composition and diversity(7–10). The analysis conducted on samples from these trials also led to the development of the Microbiome Health Index (MHI-A)—a microbiome-based biomarker of antibiotic-induced dysbiosis and restoration (11). RBX2660 treatment is associated with a significant shift of MHI-A to levels associated with those observed in healthy subjects(11).

To assess the functional impact of the microbiome changes observed with RBX2660 treatment, this study aimed to measure changes in fecal BA compositions from participants in the PUNCH CD2 Phase 2 trial. Building on a few studies that have focused on analytic rigor of fecal BA measurement (12–15), we aimed to develop a highly quantitative and sensitive assay for targeted metabolomic assessment of BAs. This method was then applied to samples from a subset of participants in the PUNCH CD2 randomized, double-blind, placebo-controlled Phase 2 trial of RBX2660 for rCDI. Results indicate that BA changes were consistent with microbiome changes and may provide clues for why RBX2660 reduced rCDI in that trial. Moreover, the resulting method may therefore be a valuable and generalized tool for rCDI diagnosis and assessing response to treatment.

## METHODS

### Preparation of BA standards

BA standards and d4 deuterated internal standards were obtained from Cayman Chemical or Steraloid (Table 1). BAs in powder form provided by the vendor were directly dissolved in methanol. Standards were pooled to create a mixture for use in the standard curve and QC samples (see below).

**Table 1.** Bile acids from Cayman Chemicals:

| **Abbreviation** | **Bile Acid Name** | **Cat#** |
| --- | --- | --- |
| T-α-MCA | Tauro-α-muricholic Acid | 20288 |
| CA | Cholic Acid | 20250 |
| CDCA | Chenodeoxycholic Acid | 10011286 |
| d4-CA | d4-Cholic Acid | 20849 |
| d4-GCDCA | d4-Glyco Chenodeoxycholic Acid | 21890 |
| d4-TCA | d4-Tauro Cholic Acid | 21891 |
| d4-DCA | d4-Deoxycholic Acid | 20851 |
| d4-LCA | d4-Lithocholic Acid | 20831 |
| d4-UDCA | Ursodeoxycholic Acid | 21892 |
| d4-GCA | Glyco Cholic Acid | 21889 |
| DCA | Deoxycholic Acid | 20756 |
| GCA | Glyco Cholic Acid | 20276 |
| GCDCA | Glyco Chenodeoxycholic Acid | 16942 |
| GDCA | Glyco Deoxycholic Acid | 20274 |
| GHCA | Glyco Hyocholic Acid | 22670 |
| GLCA | Glyco Lithocholic Acid | 21723 |
| GUDCA | Glyco Ursodeoxycholic Acid | 21698 |
| HCA | Hyocholic Acid | 20293 |
| HDCA | Hyodeoxycholic Acid | 20294 |
| LCA | Lithocholic Acid | 20253 |
| TCA | Tauro Cholic Acid | 16215 |
| TCDCA | Tauro Chenodeoxycholic Acid | 20275 |
| TDCA | Tauro Deoxycholic Acid | 15935 |
| THCA | Tauro Hyocholic Acid | 22669 |
| THDCA | Tauro Hyodeoxycholic Acid | 21956 |
| TLCA | Tauro Lithocholic Acid | 17275 |
| TUDCA | Tauro Ursodeoxycholic Acid | 20277 |
| T-β-MCA | Tauro-β-muricholic Acid | 20289 |
| UDCA | Ursodeoxycholic Acid | 15121 |
| α-MCA | α-muricholic Acid | 20291 |
| β-MCA | β-muricholic Acid | 20287 |
| ω-MCA | ω-muricholic Acid | 20292 |

**Bile acids from Steraloids:**
| **Abbreviation** | **Bile Acid Name** | **Cat#** |
| --- | --- | --- |
| 7-KLCA | 7-Ketolithocholic Acid | C1600-000 |
| 7-DHCA/7-KDCA | 7-Dehydrocholic Acid/7-ketodeoxycholic Acid | C1250-000 |
| 3-DHCA | 3-Dehydrocholic Acid | C1272-000 |
| Iso-LCA | Iso-lithocholic Acid | C1475-000 |
| 6-7-DKLCA | 6,7-Diketolithocholic Acid | C1485-000 |
| 7,12 DKLCA | 7,12-Diketolithocholic Acid | C1500-000 |
| 12-KLCA/12-KDCA | 12-Ketolithocholic Acid/12-Ketodeoxhycholic Acid | C1650-000 |
| DHLCA/3-KLCA | Dehydrolithocholic Acid/3-Ketolithocholic Acid | C1750-000 |
| DHCA | Dehydrocholic Acid | C2000-000 |
| 3-KDCA | 3-Ketodeoxycholic Acid | C1725-000 |

### Human Fecal Sample Preparation

Clinical trial participants voluntarily provided fecal samples per consent requirements (NCT02299570). The institutional review board/research ethics board (IRB/REB) at each participating center approved the study protocol. Wet fecal samples were diluted with saline to a final concentration of 0.3 g /ml. The suspension was mixed thoroughly using a mechanical homogenizer. The homogenate was gauze-filtered to remove insoluble and oversized matter if necessary, aliquoted into 96-well plates and stored in the freezer (−50C) prior to further processing.

### Sample Processing

A 20X working stock solution (in methanol) of the BA standard mixture was made and used for a 10-point calibration curve with the dynamic range of 10 – 5000 ng/ml. Another 20X working stock solution (in methanol) was prepared for quality controls and used for a 4-point curve with target concentrations of 30, 60, 300 and 3000 ng/mL. Internal standards, d4-UDCA and d4-LCA were prepared at concentration of 2500 ng/ml, whereas d4-GCDCA, d4-CA, d4-GCA, d4-TCA, and d4-DCA were prepared at 250 ng/mL in 0.1 % formic acid and methanol and mixed in equal volumes to create the internal standard mixture. This solution also served as the extraction buffer to precipitate proteins in the biological matrix.

For standard curve samples, the BA standard mixture was diluted 20X in saline and further diluted 5X in extraction buffer. A 5 µL volume was injected into liquid chromatography tandem mass spectrometry (LC-MS/MS) system for quantification.

The “blank” matrix (control stool) contains varying levels of BAs, amongst other extensive matrix related interferences, which required mathematical correction for assessment of spiked analyte concentrations. While the use of a control stool as matrix could induce further error in the assay, choosing an unrelated background matrix (like a buffer or plasma) is unfavorable.

For quality control (QC) samples, 10 µL of control stool homogenate was diluted with 180 µL saline. The mixture was spiked with 10 µL of 20X working stock solution. To extract the BAs, 800 µL of the internal standard mixture was added to the sample. For QC samples, the final concentration of BAs detected was the sum of the endogenous BAs in the control stool plus the spiked concentrations from the QC working stock solution. For blank QC samples, 10 µL of control stool homogenate was diluted with 190 µL saline. BAs were directly extracted by addition 800 µL of the internal standard mixture. The mixtures were vortexed for at least 3 minutes. After centrifugation, 200 µL of the mixture was taken and 5 µL was injected to the LCMS.

For test samples, 10 µL of fecal homogenate was diluted with 190 µL saline. BAs were directly extracted by addition of 800 µL of the internal standard mixture. The mixture was vortexed for at least 3 minutes. After centrifugation, 200 µL of the mixture was taken and 5 µL was injected into LC-MS/MS. A 20X dilution of test samples was selected so that the fecal homogenates can be easily and reliably drawn using a reasonably high volume (minimum 10 µL) to maximize pipetting accuracy.

### LC-MS/MS Analytical Method

A dual column setup was designed to maximize throughput by allowing one column to be used while the other was being regenerated.

LC Method (30 min/ sample):

- UPLC Column: Waters BEH C18 1.7 µM, 2.1 × 150 mm Column
- Mobile Phase A: 20 mM Ammonium Acetate 0.1% Formic Acid in Water
- Mobile Phase B: 20 mM Ammonium Acetate 0.1% Formic Acid in 1:3 Methanol:Acetonitrile
- Gradient: 35% B for 5 min, 35%-100% B in 20 min, 5 min re-equilibration
- Flow Rate: 0.2 ml/min

The LC was connected to a QTRAP 5500 MS/MS instrument (Sciex, Toronto, Canada). Using individual BA standards MS/MS conditions were optimized for Multiple Reaction Monitoring (MRM) detection by infusion of the BA standard. With the use of the ammonium acetate in the mobile phase, improved sensitivity and selectivity was obtained as the BAs formed ammonium adduct precursor ions(13). LCA, Iso-LCA and d4-LCA were detected in negative ion mode while all other BAs were detected in positive ion mode. The retention time of the different BAs were recorded by injected BA standards into the LC-MS/MS system and the MRM scan scheduling was optimized. The LC-MS/MS system was controlled using software Analyst 1.7.1.

Quantitative sample analysis were performed using Analyst software.

### Assay characterization

Accuracy and recovery in fecal matrix of the quality control samples were corrected with endogenous concentration of BAs of the blank matrix. The total volume of extraction was less than 1 ml to allow use of deep-well 96 well plates. Freeze-thaw stability was assessed by fully thawing the matrix at room temperature for 1 hour followed by overnight refreezing at -50C. Assay characterization using standard controls, control stool, and quality control samples included assessment of sensitivity (limit of detection and quantitation assessment), linearity (Standard Calibration dynamic range assessment), accuracy (intra- and inter-day experiment % accuracy), precision (Intra- and inter-day experiment %CV), and freeze-thaw stability.

### Application of Assay to Assess BA Composition in Samples from rCDI Patients Treated with RBX2660

To demonstrate proof-of-concept in assessing BA compositions in clinical samples, the assay was performed on fecal samples from rCDI participants from the PUNCH CD2 trial investigating RBX2660 to reduce recurrent CDI (PUNCH CD2B; NCT02299570). The institutional review board/research ethics board (IRB/REB) at each participating center approved the study protocol and clinical trial participants voluntarily provided fecal samples per consent requirements and. The clinical efficacy and safety outcome of this trial has been previously reported(16). Primary clinical efficacy was defined as absence of CDI recurrence at 8 weeks after the last study treatment. Participants were asked to provide fecal samples before study treatment (baseline) and at scheduled intervals after completion of study treatment. Participation in the sample collection phase of the trial was optional according to consent requirements. A microbiome analysis of fecal samples provided by trial participants has also been reported, in which RBX2660 treatment was associated with a significant shift in microbiome composition and diversity(8).

Stool samples for that study and a subset for this BA analysis were collected by participants at home and shipped in a freezer pack via overnight courier to Rebiotix (Minneapolis, MN). Upon receipt at Rebiotix, samples were aliquoted and frozen at -80° C with no added stabilizers until analysis. Time points were prior to receiving blinded RBX2660 or placebo study treatment (“BL,” or baseline) and 7, 30, and 60 days and 6, 12, and 24 months after blinded study treatment. A total of 113 participant stool samples from 27 participants who received RBX2660 and responded to the therapy were used in this study. An adequate number of placebo-treated samples were not available for metabolomic analysis, so they were excluded from this study.

### Statistics

A mixed effect model was fit to determine if levels of each bile acid were statistically different after treatment at timepoints 7, 30 and 60 days when compared to baseline. The repeated measures factor was accounted for by treating the subject as the random effect in the model. The model fit was bile acid as a function of the time points for each of the 8 bile acids. It was observed for each of the models that the bile acid was significantly different after treatment when compared to baseline, with p values < 0.05.

## RESULTS

### Assay principle and development

We sought to develop a multiplexed, high-throughput, and quantitative method to measure fecal BAs from clinical trial participants. Tandem liquid chromatography-mass spectrometry was used to detect abundant and rare BA species from wet fecal samples in a targeted manner using a BA standard library. Conventional primary and secondary BAs, including GCA, TCA, TCDCA, GCDCA, CA, CDCA, LCA, and DCA, were measured as well as typically rare secondary BA derivatives totaling 35 different BA species within each stool sample (**Table 1**). Throughput was increased by processing samples in a 96-well format. Additionally, the procedure was streamlined by utilizing a dual column LC system, thereby reducing the overall runtime to 30 minutes per sample (**Supplementary Figure 1**). Procedures were optimized to maximize extraction efficiency. Several controls within each experiment and each sample were embedded to control for technical variability inherent to processing fecal samples. The assay conditions used in each experiment are outlined in **Figure 1A** with representative chromatograms in **Figure 1B**. The *standard control* sample was used for absolute quantification of experimental samples. The *stool control* was a benchmark stool sample we used to develop the assay and to assess consistency throughout experiments. The *quality control* samples with spiked concentrations of each BA were used to assess extraction efficiency and to control for technical error during sample processing. *Experimental samples* were run alongside controls for each experiment. Altogether these controls allowed for normalization of data across samples and across days. Additionally, these procedures produced a high-confidence assay that is efficient, allowing for processing of hundreds of samples within a few days.

**Figure 1.**
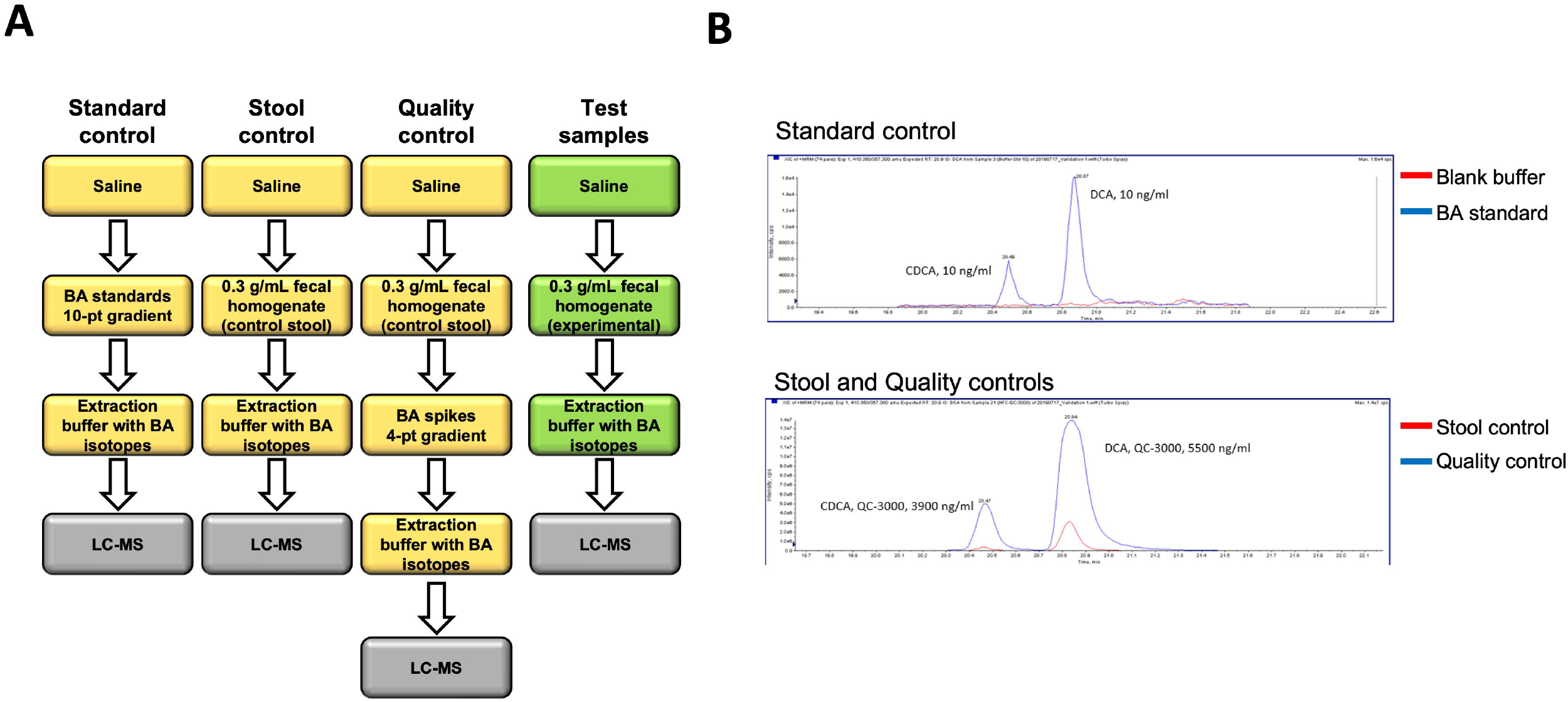
Assay procedures and readouts. **(A)** Scheme of sample preparation for controls (yellow) and test samples (green) processed in parallel. **(B)** Representative chromatograms of two standards (CDCA and DCA) in blue and a blank buffer in red (top). Representative control sample in red showing natural levels of indicated BAs and control sample with spiked BAs in blue.

### Assay properties and characterization

To evaluate assay performance, we characterized several properties including sensitivity, linearity, accuracy and precision. Limit of detection (LoD) as well as the lower limit of quantification (LoQ) and upper limit of quantification (ULoQ) of each BA were within workable range of expected amounts in human fecal matter (**Figure 2A**). The quantification range for most BA species was 10 – 5000 ng/ml highlighting the broad dynamic range of the assay. Linearity was determined by standards, with regression fitting (r^2^) typically being >0.99 for all measured BAs in multiple experiments. Intra- and inter-day assay performance were measured for 19 BAs to assess accuracy and precision. Assay precision (%CV) largely ranged within 5-20% and accuracy (% accuracy) ranged within 75-125% across experiments performed on 3 separate days (**Figure 2B**). These data indicate that our method is robust, precise, accurate, and reproducible.

**Figure 2.**
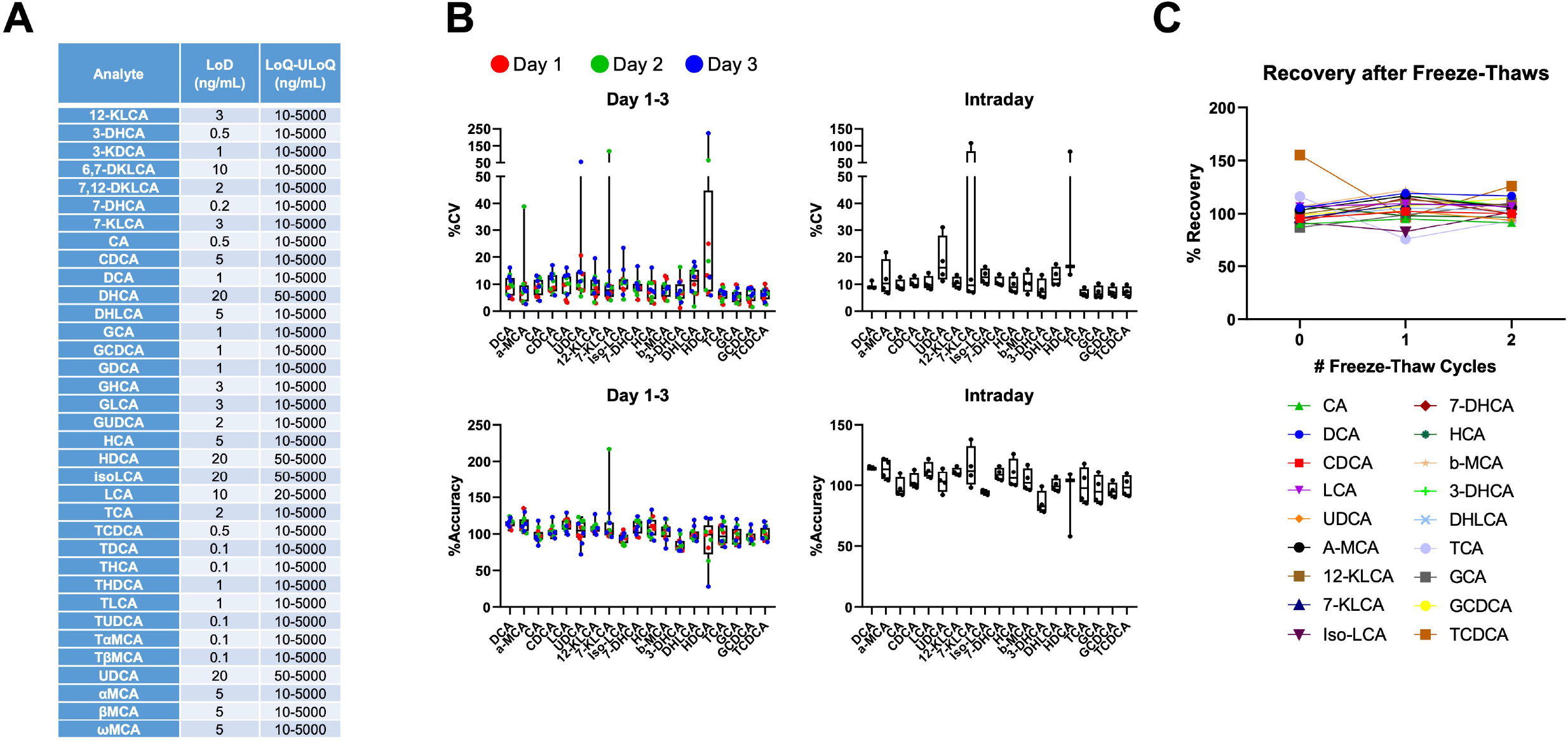
Assay quantification and sensitivity in fecal matrix. **(A)** Limit of detection (LoD) and limit of quantification (LoQ) to upper limit of quantification (ULoQ) of the indicated BA highlighting the sensitivity of our method. **(B)** Inter- and Intra-day runs show high precision (%CV) and high accuracy of quality control samples. 3 independent experiments were run with each BA using a 4-point concentration gradient. **(C)** Detection of fecal BA after 0, 1, and 2 rounds of freeze-thaws.

Since clinical samples are collected in different locations and kept frozen for storage, samples must be thawed at least once to allow for processing. Using the control stool sample, we performed freeze-thaw cycling and measured the stability of 16 representative BAs. After 2 rounds of freeze-thawing, the concentration of all measured BAs did not significantly change (**Figure 2C**). Notably, the clinical samples were not expected to undergo more than 2 freeze-thaw cycles during the processing steps so stability was not measured beyond 2 cycles. Taken together, these data suggest that BAs are relatively stable in fecal matter and confirm that our procedures are well-optimized for the analysis of clinical samples.

### Proof-of-Concept of BA Metabolic Assessment in Clinical Samples

To test our approach, we measured BAs from available fecal samples of participants in the PUNCH CD2 trial of RBX2660 for rCDI. Total BA levels were elevated at baseline and moderately reduced after study treatment, remaining stable throughout the follow-up period (Day 7 – 720) (**Supplementary Figure 2A**). Primary BAs were the dominant BA species at baseline, consistent with the expected loss of commensals associated with broad-spectrum antibiotic treatment (**Figure 3**). As early as 7 days after RBX2660 treatment, there was a significant drop in primary BAs concurrent with increased secondary BAs, and this profile was sustained through 24 months after treatment. Over the entire time course, the most abundant BAs detected in our sample set were primary BAs that are synthesized in the liver (GCA, TCA, GCDCA, TCDCA, CA, CDCA) and the secondary BAs (LCA, DCA) that typically dominate the fecal BA pool in healthy individuals(17,18). Consistent with the patterns of LCA and DCA levels, other 7-dehydroxylated secondary BA (12-KLCA, DHLCA, isoLCA, and 3-KDCA) were also lacking in baseline patients and increased over the study period. Overall, these data confirmed that our method produces results that are consistent with previous reports that have measured fecal BA levels in rCDI patients.

**Figure 3.**
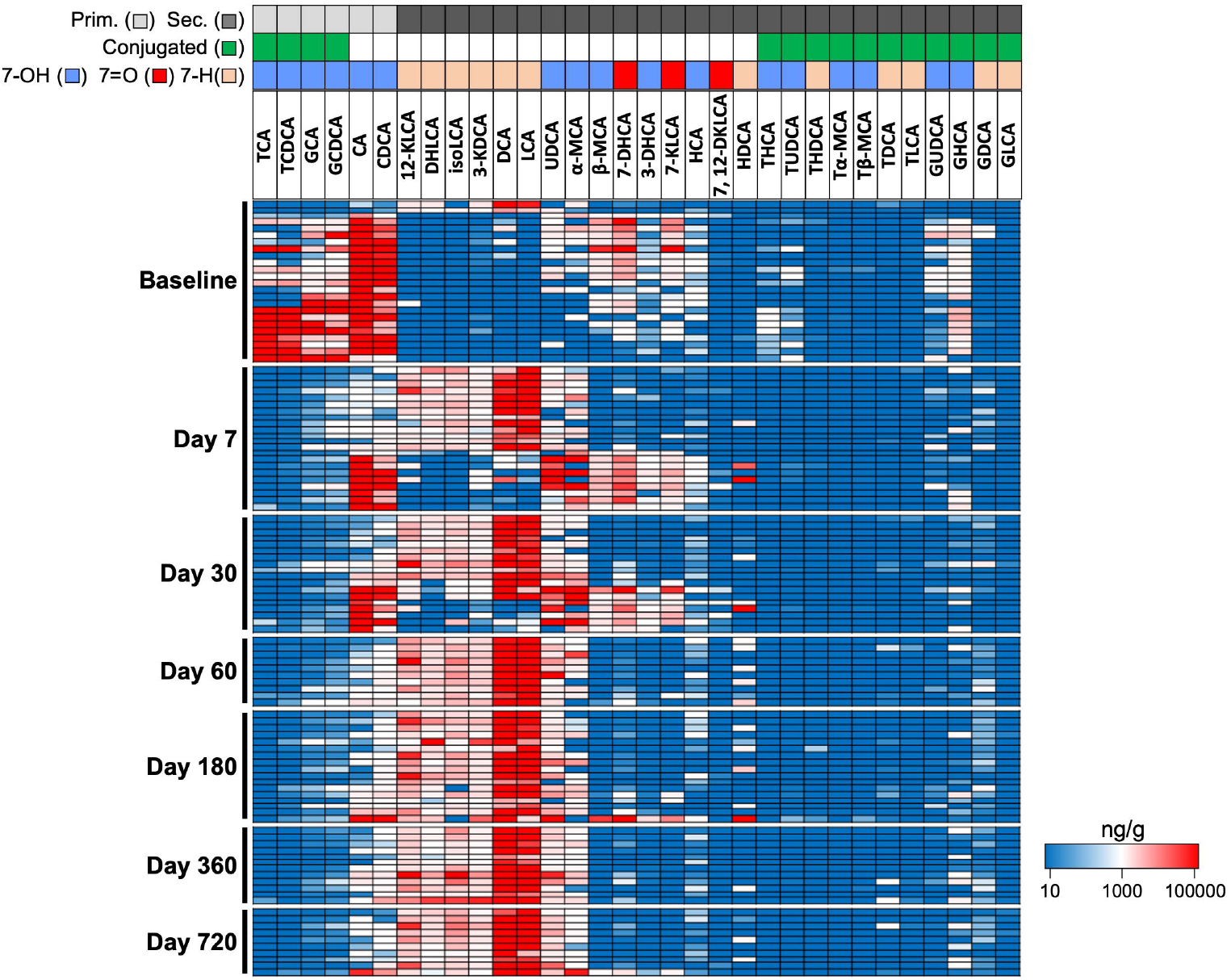
Large-scale BA changes detected in rCDI patients treated with RBX2660. Heatmap of fecal BA levels (ng/g) in wet fecal matter from PUNCH CD2 trial participants. Each row is a single patient sample, with rows grouped by timepoints (at or days post-baseline) of when the stool samples were collected. Annotations in boxes above the heatmap include primary (light gray) and secondary (dark gray) BA, conjugate (green) and deconjugated (white) BA, as well as 7-hydroxylation (7-OH, blue), 7-oxo (7=O, red), and 7-dehydroxylated (7-H, yellow) BA.

Our method allowed us to measure up to 35 BAs in a targeted manner including non-standard or typically rare secondary BA species that contain a 7-hydroxyl or 7-oxo group (UDCA, a-MCA, b-MCA, 7-KLCA, HCA, 7DHCA, and 3-DHCA). Interestingly, this group of secondary BAs were moderately elevated in many baseline patient samples representing up to 50% of the total fecal BA pool (**Supplementary Figure 2B**). They appeared in conjunction with deconjugated CDCA and CA in baseline patients, a trend that was also apparent in day 7 and day 30 samples. In lieu of commensals that normally carry out 7-dehydroxylation to form DCA and LCA, it is conceivable that CDCA and CA can be substrates for the formation of these typically rare secondary BAs in baseline patients. Indeed, 7-hydroxyl and 7-oxo secondary BAs are chemically differentiated by single biochemical reactions that are typically carried out by microbiota(19) (**Supplementary Figure 3**).

An analysis was made to assess whether there are meaningful changes over time to key bile acids that have been shown to regulate the *C. difficile* lifecycle. TCA and its closely related species CA and GCA are well-studied germinants of *C. difficile* whereas LCA and DCA are associated with suppression of *C. difficile* outgrowth(6,20–22). TCA levels were highly elevated in baseline samples and precipitously dropped as early as one week post RBX2660 treatment (**Figure 4A**). A similar pattern was detected for CA and GCA. Conversely, LCA and DCA levels were relatively low at baseline and rapidly increased several orders of magnitude after RBX2660 treatment (**Figure 4B**). Despite the low sample numbers, each BA at baseline was significantly different after treatment (p < 0.05). Taken together, the changes to these specific BA show that there are functional changes to the microbiome occurring post-baseline.

**Figure 4:**
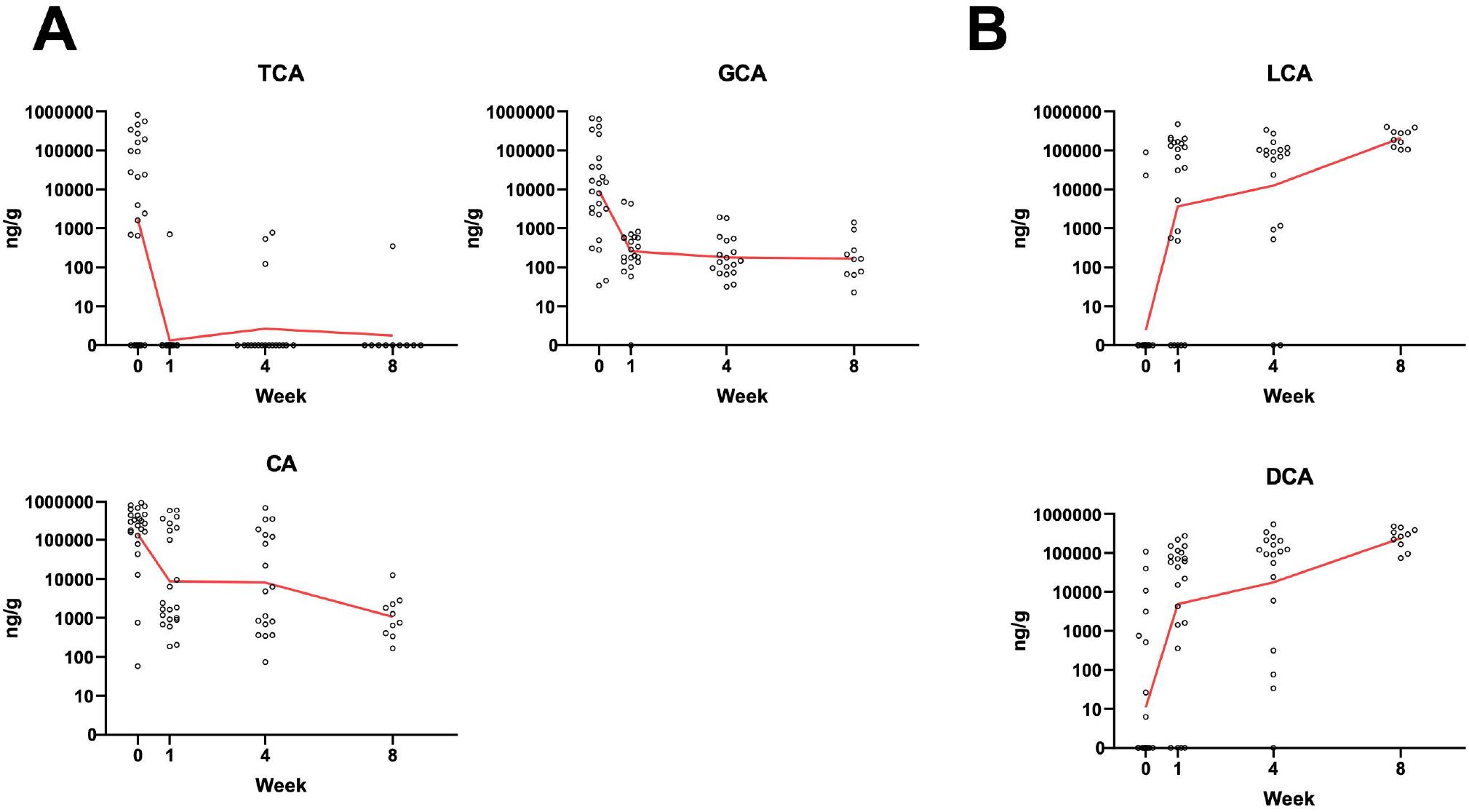
Quantitative and specific BA changes. **(A)** Absolute fecal levels of the primary Bas TCA, GCA, CA known to promote *C. diff* germination. **(B)** Absolute fecal levels of the secondary BAs LCA and DCA associated with suppression of *C. diff* outgrowth. Samples from study participants were taken at baseline, day 7, day 30, and day 60 post RBX2660 treatment. Red line represents the geometric mean. Statistical significant was determined by a linear mixed model and the changes from baseline were significantly different for all BAs (p<0.05).

The overall impact of BAs on the *C. difficile* lifecycle is complex, but in general it is thought that a fecal BA composition enriched for primary BAs is associated with dysbiosis and therefore is associated with an increased risk of CDI recurrence. In contrast, a fecal BA profile dominated by secondary BAs is normally seen in the healthy gut, which is resistant to *C. difficile* outbreaks(18). To quantify mean changes to BA compositions in the study population and to compare to after treatment, we grouped all detected BAs into four categories including primary conjugated, primary deconjugated, secondary deconjugated, and secondary conjugated. The means of each group represented on a fractional graph confirmed that primary BAs dominated baseline samples but were efficiently replaced by secondary BAs as early as 7 days post RBX2660 treatment. Secondary BA levels progressively increased over time plateauing after about 8 weeks post RBX2660 treatment (**Figure 5**). Taken together, our results show that BA metabolism is profoundly altered from before to after RBX2660 administration, shifting towards a profile that is expected to have higher resistance to *C. difficile* colonization.

**Figure 5:**
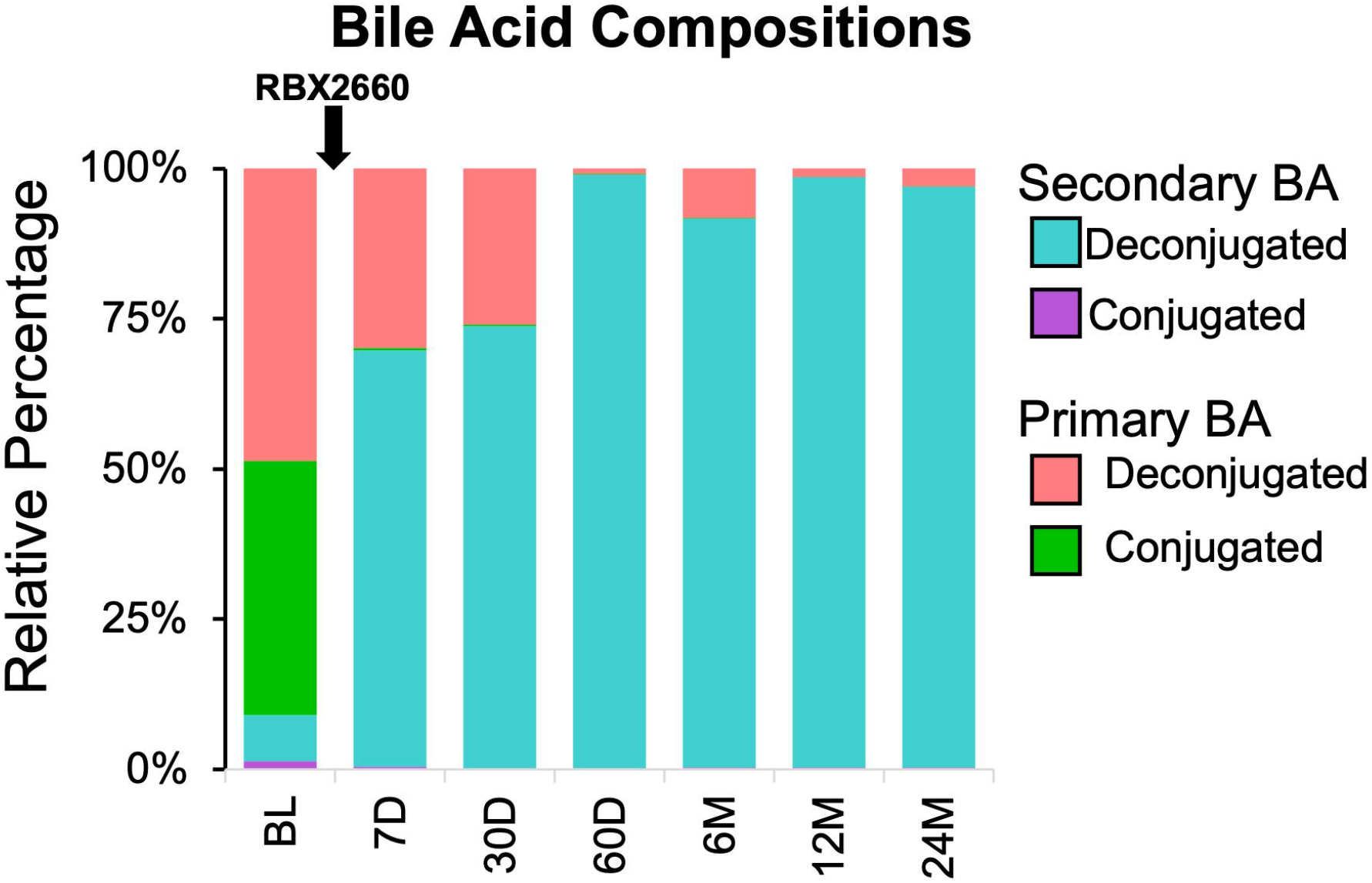
BA compositions over time. All detected bile acids were categorized and grouped as either primary or secondary and conjugated or deconjugated to highlight the average changes to fecal BA composition over the course of the study in rCDI patients receiving RBX2660.

## DISCUSSION

Here, we describe the development and characterization of a bioanalytical method for targeted metabolomic assessment of BAs in human fecal samples. The assay involved quantification of up to 35 BAs in feces at varying dynamic ranges, designed according to the anticipated levels in disease and in health. With a focus on reproducibility and throughput, we optimized a protocol that is streamlined and well-controlled allowing for multiplexed assessment of BAs in hundreds of fecal samples across different experiments. Using this assay, we showed that large-scale fecal BA changes occurred in rCDI trial participants that received RBX2660. The observed high relative abundance of primary BAs at baseline is consistent with the expected consequence of broad-spectrum antibiotic treatment(23,24). The observed shift towards a secondary BA dominant profile in post-baseline samples is consistent with previous findings in rCDI patients receiving FMT(6,25–27). Our results demonstrate that this method is rigorous and are in line with previous observations in rCDI patients.

The accuracy and precision of the assay were studied across 3 assay days and found to be highly reproducible. The assay also indicated that BAs are stable up to 2 freeze-thaw cycles (for the 16 analytes analyzed), so samples from human trials should be analyzed within 2 freeze-thaw cycles. Room temperature stability of the fecal samples was not assessed, although a previous study has shown that temperature is not a confounding factor(12). In addition, the assay was not conducted under GLP conditions, but the characterization procedures are based on common industry standards for assessment of quality and reproducibility of LC-MS based bioanalytical methods(28,29).

Water content in feces can vary from 60-90%, thus potentially complicating absolute fecal biomarker measurements(30,31). However, lyophilizing fecal matter prior to BA extraction can lead to loss of some BAs, a confounding factor that would need to be corrected for by internal standards(12). Therefore, to reduce procedural burden and technical error, we extracted BAs from wet feces and designed an efficient extraction protocol to streamline processing time. Notably, the omission of a lyophilization step may contribute to the high sensitivity of the method, with many BAs detected in the low nM range in this study. Our use of internal deuterated standards together with wet fecal matter further controlled for technical variation enabling us to measure absolute levels of fecal BAs across hundreds of samples. Notably, the consistency of total BA levels across our study samples (**Supp Fig 2A**) highlight that water content is not a major source of variation in the measurement of absolute fecal BA levels in our study population.

The unique longitudinal design of our clinical trial enabled us to track fecal BA profile in participants over a 2-year period in some cases. We found that there is an immediate deconjugation of primary bile acids as early as 7 days post-baseline, yielding CDCA and CA that can serve as substrates for the synthesis of secondary BAs. When assessing specific BA abundance over the efficacy period of the trial (8 weeks post-RBX2660 treatment), specific BA species such as the *C. difficile* germinant TCA were among these showing a sustained drop in levels (p <0.05). Reciprocally, secondary bile acids such as LCA and DCA, associated with *C. difficile* growth suppression, significantly increased (p<0.05). By 8 weeks post-baseline, the fecal BA profile was dominated by the secondary BAs LCA and DCA. Notably, this shift in microbiome-dependent BA metabolism was sustained through at least 720 days. Acknowledging the low sample numbers and lack of head-to-head data, secondary BA appeared to restore more efficiently after RBX2660 administration in this trial than after antibiotics alone, including some in late-stage clinical development(32,33).

In addition to the conventional BAs that we and others have studied, our assay detected a wide range of secondary BAs that are expected to be uncommon in healthy human feces. Some of which have previously not been measured in a targeted manner in CDI patients. We found that a subset of baseline patients had elevated levels of these otherwise rare secondary BAs (UDCA, aMCA, bMCA, 7-KLCA, HCA, 7-DHCA, and 3-DHCA). These secondary BAs were abundant representing on average up to 20% (3.4 - 50% range) of the total fecal BA pool in a subset of baseline patients. The emergence of these BAs also appeared anti-correlated with conjugated primary BAs, suggesting that deconjugation of primary BAs occurs prior to their biochemical conversion to secondary BAs, as is the case for the synthesis of LCA and DCA(4,34). Notably, these typically rare secondary BAs contained either a 7-hydroxyl or 7-oxo group indicating that the microbiota capable of carrying out 7-dehydroxylation was lacking in baseline patients. The emergence of LCA and DCA and other 7-dehyroxylated derivatives (12-KLCA, DHLCA, isoLCA, 3-KDCA) starting on day 7 post-baseline revealed that commensals with 7-dehydroxylating activity were being restored over time in rCDI patients after RBX2660 treatment. Consistent with this, the restoration of secondary BAs post-baseline coincided with a shift to a healthier MHI-A in the same cohort(8,11). Notably, a healthier MHI-A is partially defined by increased relative operational taxonomic units (OTUs) from the *Clostridia* class, which is known to harbor critical commensals that express 7-dehydroxylating genes(3–5).

In patient samples from the PUNCH CD2 trial, we demonstrated reduced fecal levels of primary BAs and restored secondary BAs in patients who responded to treatment with RBX2660 (*i*.*e*. no recurrence of CDI in 8 weeks post treatment). A more comprehensive analysis, including comparing RBX2660 to placebo administration, will be conducted in a future study of a large phase 3 trial of RBX2660. Combining traditional metagenomic sequencing with this robust bioanalytical method for targeted quantification of BAs will allow for a deeper analysis that not only examines restoration of microbiome diversity after treatment with RBX2660 but also assesses functional microbial activity that mechanistically contributes to the observed reduction in recurrent CDI.

## Data Availability

The datasets presented in this article are not readily available because they are restricted by an ongoing commercialization process. Requests to access the datasets should be directed to KB.

## ACKNOWLEDGEMENTS

The authors would like to acknowledge the support of nurses, administration, staff, and investigators for all clinical trial sites; and the manuscript assistance of Amy Moore, PhD from Moore BioTranslation. We thank Christella Widjaja for assistance with data auditing.

## AUTHOR CONTRIBUTIONS

RP, NF, KS, KB and BCF performed study concept and design; RP, NF, KS, KB and BCF performed development of methodology and writing, review and revision of the paper; RP, NF, KS, KB and BCF provided acquisition of the data; CG and BS provided statistical analysis; and, all authors provided analysis and interpretation of data. All authors read and approved the final paper.

## ETHICS APPROVAL AND CONSENT TO PARTICIPATE

The institutional review board/research ethics board (IRB/REB) at each participating center approved the study protocol and informed consent form (Quorum Review, University of Chicago Biological Sciences Division/University of Chicago Medical Center; Western IRB; Louis Stokes Cleveland Department of Veterans Affairs Medical Center IRB; Henry Ford Health System IRB; University of Pennsylvania Institutional Review Board; St. Vincent’s Hospital and Health Care Center, Inc IRB; Loyola University Chicago Health Sciences Division Institutional Review Board for the Protection of Human Subjects; Mayo Clinic Institutional Review Boards; Weill Cornell Medical College IRB; IRB of St. Rita’s Health Partners; HealthPartners Institute for Education and Research IRB; Hamilton Integrated Research Ethics Board; University of British Columbia Clinical Research Ethics Board; Washington University Human Research Protection Office). All study participants provided written informed consent. An independent medical monitor provided safety oversight. All methods were carried out in accordance with relevant guidelines and regulations.

## REFERENCES

1. Sheitoyan-Pesant C, Abou Chakra CN, Pépin J, Marcil-Héguy A, Nault V, Valiquette L. Clinical and Healthcare Burden of Multiple Recurrences of Clostridium difficile Infection. Clin Infect Dis. 2016 Mar 1;62(5):574–80.

2. Mullish BH, Allegretti JR. The contribution of bile acid metabolism to the pathogenesis of Clostridioides difficile infection. Therap Adv Gastroenterol. 2021 May 28;14:17562848211017724.

3. Reed AD, Theriot CM. Contribution of Inhibitory Metabolites and Competition for Nutrients to Colonization Resistance against Clostridioides difficile by Commensal Clostridium. Microorganisms [Internet]. 2021 Feb 12;9(2). Available from: http://dx.doi.org/10.3390/microorganisms9020371

4. Ridlon JM, Kang D-J, Hylemon PB. Bile salt biotransformations by human intestinal bacteria. J Lipid Res. 2006 Feb;47(2):241–59.

5. Buffie CG, Bucci V, Stein RR, McKenney PT, Ling L, Gobourne A, et al. Precision microbiome reconstitution restores bile acid mediated resistance to Clostridium difficile. Nature. 2015 Jan 8;517(7533):205–8.

6. Weingarden AR, Dosa PI, DeWinter E, Steer CJ, Shaughnessy MK, Johnson JR, et al. Changes in Colonic Bile Acid Composition following Fecal Microbiota Transplantation Are Sufficient to Control Clostridium difficile Germination and Growth. PLoS One. 2016 Jan 20;11(1):e0147210.

7. Orenstein R, Dubberke ER, Khanna S, Lee CH, Yoho D, Johnson S, et al. Durable reduction of Clostridioides difficile infection recurrence and microbiome restoration after treatment with RBX2660: results from an open-label phase 2 clinical trial. BMC Infect Dis. 2022 Mar 12;22(1):245.

8. Blount KF, Shannon WD, Deych E, Jones C. Restoration of Bacterial Microbiome Composition and Diversity Among Treatment Responders in a Phase 2 Trial of RBX2660: An Investigational Microbiome Restoration Therapeutic. Open Forum Infect Dis. 2019 Apr;6(4):ofz095.

9. Langdon A, Schwartz DJ, Bulow C, Sun X, Hink T, Reske KA, et al. Microbiota restoration reduces antibiotic-resistant bacteria gut colonization in patients with recurrent Clostridioides difficile infection from the open-label PUNCH CD study. Genome Med. 2021 Feb 16;13(1):28.

10. Kwak S, Choi J, Hink T, Reske KA, Blount K, Jones C, et al. Impact of investigational microbiota therapeutic RBX2660 on the gut microbiome and resistome revealed by a placebo-controlled clinical trial. Microbiome. 2020 Aug 31;8(1):125.

11. Blount K, Jones C, Walsh D, Gonzalez C, Shannon WD. Development and Validation of a Novel Microbiome-Based Biomarker of Post-antibiotic Dysbiosis and Subsequent Restoration. Front Microbiol. 2021;12:781275.

12. Shafaei A, Rees J, Christophersen CT, Devine A, Broadhurst D, Boyce MC. Extraction and quantitative determination of bile acids in feces. Anal Chim Acta. 2021 Mar 15;1150:338224.

13. Wegner K, Just S, Gau L, Mueller H, Gérard P, Lepage P, et al. Rapid analysis of bile acids in different biological matrices using LC-ESI-MS/MS for the investigation of bile acid transformation by mammalian gut bacteria. Anal Bioanal Chem. 2017 Feb;409(5):1231–45.

14. Perwaiz S, Mignault D, Tuchweber B, Yousef IM. Rapid and improved method for the determination of bile acids in human feces using MS. Lipids. 2002 Nov;37(11):1093–100.

15. Liu Y, Rong Z, Xiang D, Zhang C, Liu D. Detection technologies and metabolic profiling of bile acids: a comprehensive review. Lipids Health Dis. 2018 May 23;17(1):121.

16. Dubberke ER, Lee CH, Orenstein R, Khanna S, Hecht G, Gerding DN. Results From a Randomized, Placebo-Controlled Clinical Trial of a RBX2660-A Microbiota-Based Drug for the Prevention of Recurrent Clostridium difficile Infection. Clin Infect Dis. 2018 Sep 28;67(8):1198–204.

17. Theriot CM, Koenigsknecht MJ, Carlson PE Jr, Hatton GE, Nelson AM, Li B, et al. Antibiotic-induced shifts in the mouse gut microbiome and metabolome increase susceptibility to Clostridium difficile infection. Nat Commun. 2014;5:3114.

18. Winston JA, Theriot CM. Impact of microbial derived secondary bile acids on colonization resistance against Clostridium difficile in the gastrointestinal tract. Anaerobe. 2016 Oct;41:44–50.

19. Guzior DV, Quinn RA. Review: microbial transformations of human bile acids. Microbiome. 2021 Jun 14;9(1):140.

20. Sorg JA, Sonenshein AL. Bile salts and glycine as cogerminants for Clostridium difficile spores. J Bacteriol. 2008 Apr;190(7):2505–12.

21. Sorg JA, Sonenshein AL. Chenodeoxycholate is an inhibitor of Clostridium difficile spore germination. J Bacteriol. 2009 Feb;191(3):1115–7.

22. Wilson KH. Efficiency of various bile salt preparations for stimulation of Clostridium difficile spore germination. J Clin Microbiol. 1983 Oct;18(4):1017–9.

23. Reijnders D, Goossens GH, Hermes GDA, Neis EPJG, van der Beek CM, Most J, et al. Effects of Gut Microbiota Manipulation by Antibiotics on Host Metabolism in Obese Humans: A Randomized Double-Blind Placebo-Controlled Trial. Cell Metab. 2016 Jul 12;24(1):63–74.

24. Vrieze A, Out C, Fuentes S, Jonker L, Reuling I, Kootte RS, et al. Impact of oral vancomycin on gut microbiota, bile acid metabolism, and insulin sensitivity. J Hepatol. 2014 Apr;60(4):824–31.

25. Allegretti JR, Kearney S, Li N, Bogart E, Bullock K, Gerber GK, et al. Recurrent Clostridium difficile infection associates with distinct bile acid and microbiome profiles. Aliment Pharmacol Ther. 2016 Jun;43(11):1142–53.

26. Brown JR-M, Flemer B, Joyce SA, Zulquernain A, Sheehan D, Shanahan F, et al. Changes in microbiota composition, bile and fatty acid metabolism, in successful faecal microbiota transplantation for Clostridioides difficile infection. BMC Gastroenterol. 2018 Aug 28;18(1):131.

27. Seekatz AM, Theriot CM, Rao K, Chang Y-M, Freeman AE, Kao JY, et al. Restoration of short chain fatty acid and bile acid metabolism following fecal microbiota transplantation in patients with recurrent Clostridium difficile infection. Anaerobe. 2018 Oct;53:64–73.

28. Piccoli SP, Garofolo F. Biomarker assay validation. Bioanalysis. 2018 Jun 1;10(12):889–91.

29. Center for Drug Evaluation, Research. Bioanalytical method validation guidance for industry [Internet]. U.S. Food and Drug Administration. 2020 [cited 2022 May 9]. Available from: https://www.fda.gov/regulatory-information/search-fda-guidance-documents/bioanalytical-method-validation-guidance-industry

30. Rose C, Parker A, Jefferson B, Cartmell E. The Characterization of Feces and Urine: A Review of the Literature to Inform Advanced Treatment Technology. Crit Rev Environ Sci Technol. 2015 Sep 2;45(17):1827–79.

31. Caroff DA, Edelstein PH, Hamilton K, Pegues DA, CDC Prevention Epicenters Program. The Bristol stool scale and its relationship to Clostridium difficile infection. J Clin Microbiol. 2014 Sep;52(9):3437–9.

32. Qian X, Yanagi K, Kane AV, Alden N, Lei M, Snydman DR, et al. Ridinilazole, a narrow spectrum antibiotic for treatment of Clostridioides difficile infection, enhances preservation of microbiota-dependent bile acids. Am J Physiol Gastrointest Liver Physiol. 2020 Aug 1;319(2):G227–37.

33. Garey KW, McPherson J, Dinh AQ, Hu C, Jo J, Wang W, et al. Efficacy, Safety, Pharmacokinetics, and Microbiome Changes of Ibezapolstat in Adults with Clostridioides difficile Infection: A Phase 2a Multicenter Clinical Trial. Clin Infect Dis [Internet]. 2022 Feb 4; Available from: http://dx.doi.org/10.1093/cid/ciac096

34. Batta AK, Salen G, Arora R, Shefer S, Batta M, Person A. Side chain conjugation prevents bacterial 7-dehydroxylation of bile acids. J Biol Chem. 1990 Jul 5;265(19):10925–8.

