## Supplementary Information for "HUMAN FECAL BILE ACID ANALYSIS AFTER INVESTIGATIONAL MICROBIOTA-BASED LIVE BIOTHERAPEUTIC DELIVERY FOR RECURRENT *CLOSTRIDIOIDES DIFFICILE* INFECTION"

**Supplementary Figure 1: Performance of dual column setup.** Representative MRM chromatograms of BA standards run in parallel using our dual column set up. Near identical readouts and column performance was seen throughout study. Zoomed image of a 16 min window of the run to highlight high retention time, reproducibility, and separation fidelity.

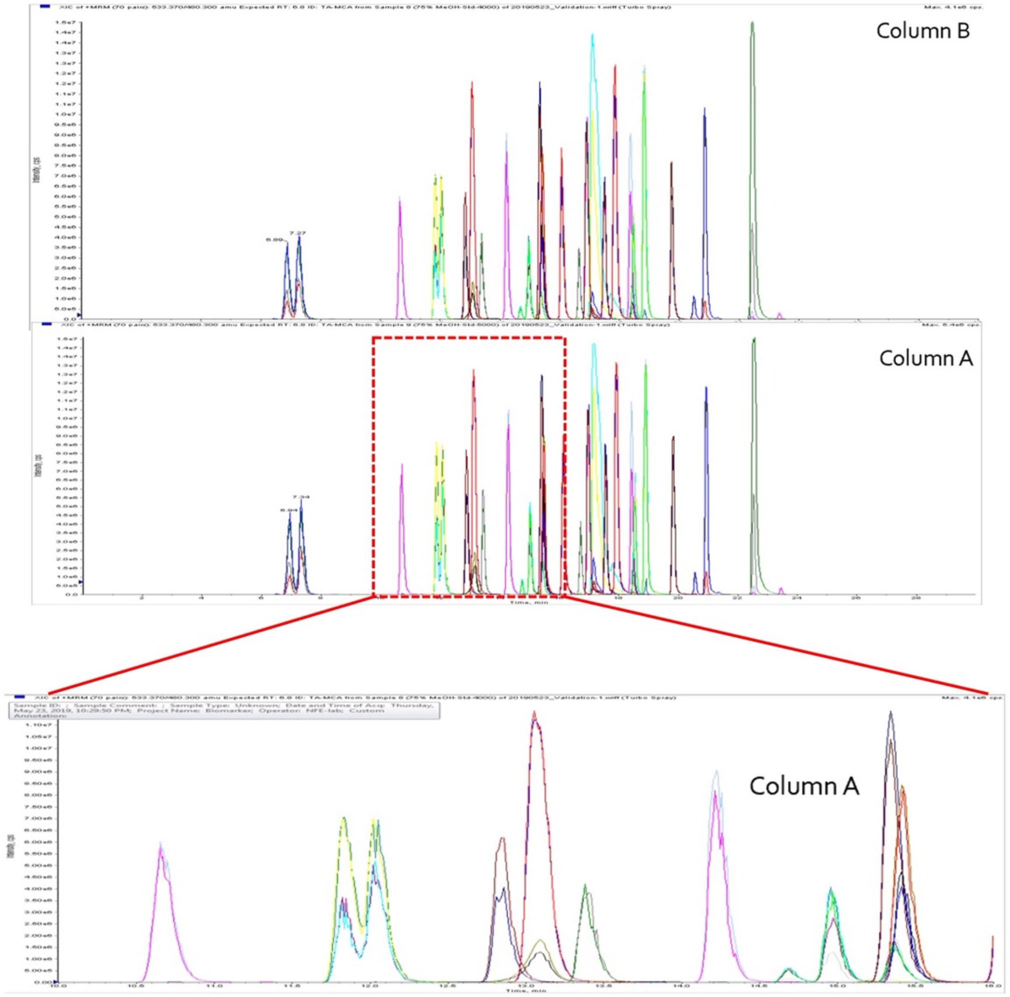

**Supplementary Figure 2: Fecal BA abundance and composition (A)**Total fecal BA levels of 35 detected BA (ng BA/g wet fecal matter) from participant samples. **(B)** Percent abundance of primary and secondary BA of baseline patients (each row) broken down by conjugation status and dehydroxylation (7-H), hydroxylation (7-OH), or oxidation (7=O) of the 7-carbon.

**A**

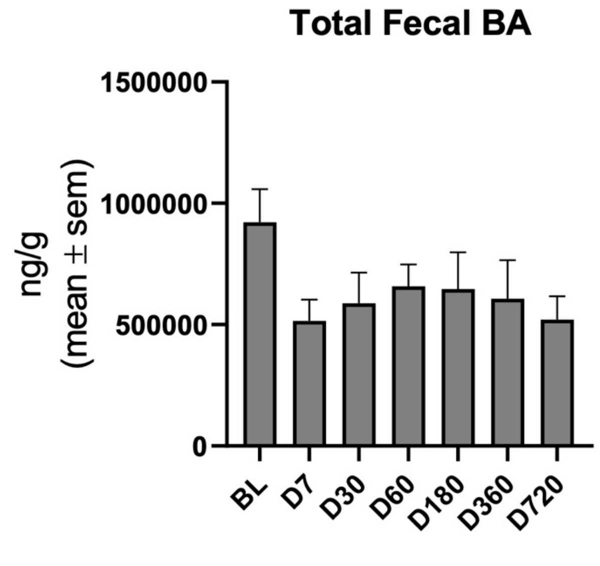


**B**

**Relative BA abundance (%) in baseline rCDI patients (n=24)**

| # | 1° Conj | 1° Unconj | 2° Unconj 7-H | 2° Unconj 7-OH /7=O | 2° Conj |
|---|---|---|---|---|---|
| 1 | 0.1 | 0.4 | 95.6 | 3.7 | 0.2 |
| 2 | 0.1 | 0.5 | 90.8 | 8.6 | 0.0 |
| 3 | 2.0 | 47.5 | 0.1 | 50.0 | 0.4 |
| 4 | 10.4 | 49.9 | 0.1 | 38.7 | 1.0 |
| 5 | 3.9 | 64.4 | 1.8 | 27.7 | 2.2 |
| 6 | 29.6 | 45.0 | 0.0 | 15.7 | 9.6 |
| 7 | 1.0 | 81.9 | 1.0 | 15.9 | 0.2 |
| 8 | 35.7 | 47.5 | 0.0 | 16.5 | 0.4 |
| 9 | 0.7 | 87.5 | 0.2 | 10.5 | 1.1 |
| 10 | 1.2 | 87.8 | 0.0 | 9.9 | 1.1 |
| 11 | 10.4 | 78.8 | 0.0 | 9.2 | 1.6 |
| 12 | 2.4 | 88.3 | 0.0 | 8.4 | 1.0 |
| 13 | 26.6 | 67.5 | 0.0 | 3.5 | 2.4 |
| 14 | 1.0 | 94.5 | 0.0 | 3.4 | 1.1 |
| 15 | 15.4 | 80.4 | 0.0 | 1.0 | 3.2 |
| 16 | 78.7 | 18.0 | 0.2 | 3.1 | 0.0 |
| 17 | 74.9 | 22.1 | 0.0 | 1.2 | 1.8 |
| 18 | 27.8 | 69.6 | 0.0 | 0.3 | 2.3 |
| 19 | 87.0 | 11.1 | 0.0 | 0.7 | 1.2 |
| 20 | 91.5 | 6.9 | 0.0 | 0.3 | 1.3 |
| 21 | 15.4 | 83.4 | 0.0 | 0.5 | 0.8 |
| 22 | 33.1 | 65.7 | 0.0 | 0.3 | 0.9 |
| 23 | 28.9 | 70.2 | 0.0 | 0.1 | 0.8 |
| 24 | 98.5 | 0.9 | 0.0 | 0.0 | 0.6 |

**Supplementary Figure 3: Enriched secondary BAs in baseline patients are separated by single biochemical steps.** Secondary BA enriched in baseline patients are separated by single biochemical steps such as epimerization, dehydroxylation, and dehydrogenation that can be carried out by microbiota. BAs in red are elevated in some baseline rCDI patients.

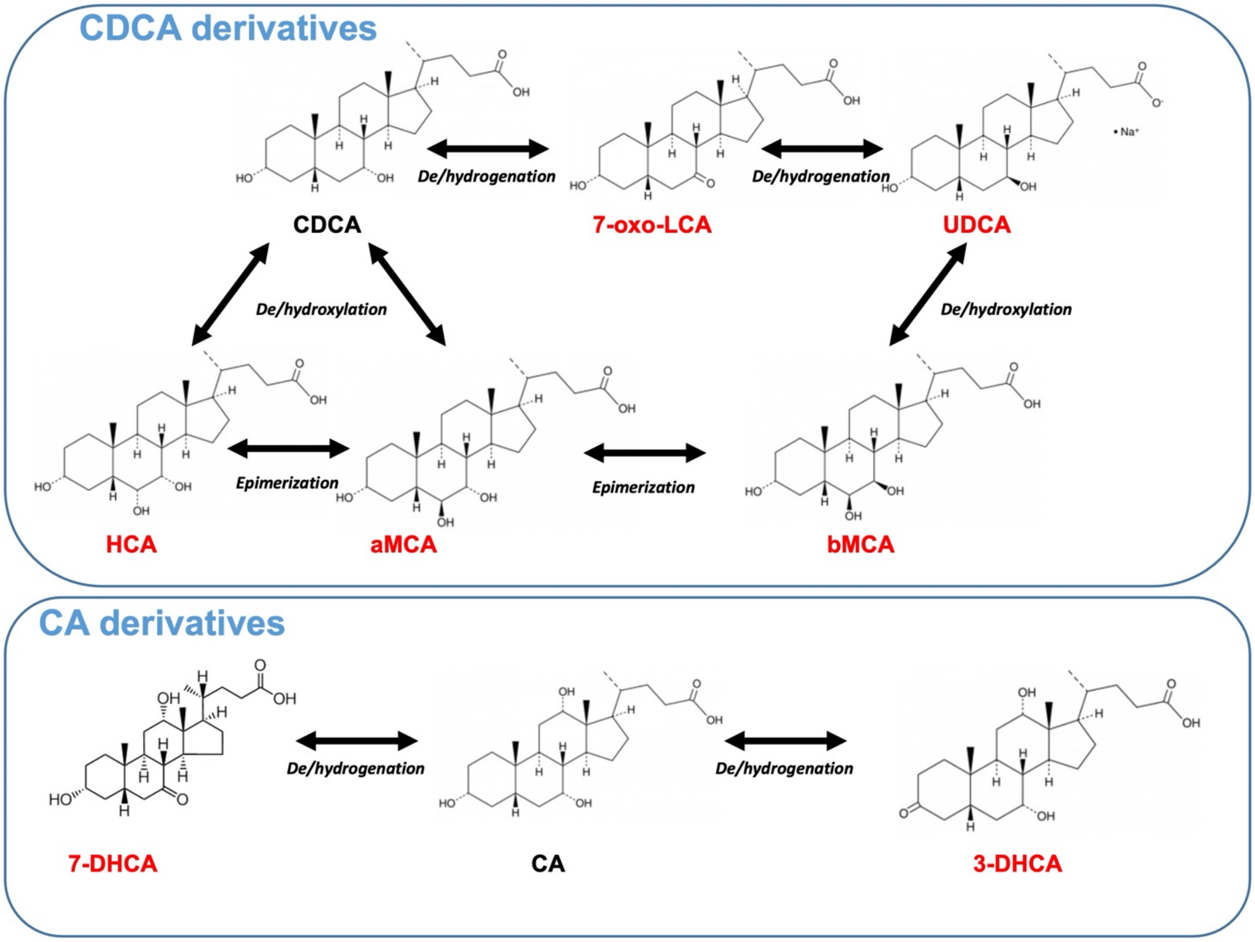
